# Are men dying more than women by COVID-19?

**DOI:** 10.1101/2020.07.06.20147629

**Authors:** Thaise Pinto de Melo, Delvan Alves Silva, Alexandre Naime Barbosa

## Abstract

We aimed to clarify if the infection and death rate by COVID-19 differ among gender in the top 50 countries with the highest death rates. Also, we investigated if secondary variables such as HDI, number of hospital beds, average age, temperature, percentage of elderly, smoker and obesity are contributing to the variability observed among countries. Meta-analyses and meta-regressions approaches were applied to official public data reported by the Word Health Organization and governments until May, 2020. A random effect model was used for the meta-analysis and heterogeneity was calculated by I^2^ statistic. There was not significative difference between men and women to be infected by COVID-19 (*P* = 0.42), though a significative difference was observed for death rate (*P* < 0.0001). High heterogeneity was observed among countries. For both infection and death rates this variability was mainly explained by the HDI (42.3% and 54.2%), average age (40.9% and 40.3%) and temperature (30.1% and 39.3%). Man are dying more than women around the word by COVID-19. Countries with highest HDI present less difference between sexes. These results reinforce that public politics promoting social isolation, health care and general well-being of the population are key factors in combating COVID-19.

## 1. Introduction

Severe acute respiratory syndrome coronavirus 2, shortened as SARS-CoV-2, is an RNA virus from coronaviruses family, as SARS-CoV that causes the recent outbreak of novel Coronavirus disease (COVID-19). In the last months we have seen a huge increasing of infections and deaths around all the word caused by this virus, which culminate in a declaration of Word Health Organization (WHO) classifying the situation as a pandemic in March of 2020 (Daga et al., 2019). According to WHO (https://www.who.int) until May, 29 of 2020 almost 5,8 million of COVID-19 cases and more than 362 thousand of deaths were registered.

Several studies have been pointed that the number of infected and deaths by Covid-19 may vary according to the gender (Cai, 2020), the age (Shi et al., 2020) and other factors as pre-existing diseases, as diabetes (Guo et al., 2020), hypertension (Fang, Karakiulakis, & Roth, 2020), chronic respiratory diseases (Shi et al., 2020), among others, also known as the risk group. In addition, the temperature also may affect the rate of respiratory infections (Mäkinen et al., 2009).

As the social isolation sum to good hygienic conditions have been recommended by the WHO as the most efficient way in combating the COVID-19, economical and infrastructure information such as the human development index (HDI) and the number of hospital beds of the country could also influence the chance of infection and death.

Thus, in the current scenario is important to have a look in what the statistics around the world are saying about the profile of infected and dead coronavirus patients. This understanding can be achieved by a compilation of the public data published from each country in the word, giving a global profile of the actual situation.

Some statistics are pointing that men present more chance to die than women, but how of this information is true around the word, i.e., this would be a reality in all countries? It would be true for the chance of infection? And which secondary factors may be affecting this? To answer these questions, we applied meta-analysis and meta-regression approaches in public data from a total of 50 countries that presented the highest death rate by COVID-19.

## 2. Material and Methods

### Data and Study Design

We applied meta-analysis and meta-regression approaches to verify differences between genders regarding the infection and death rates around the word. Two meta-analysis were performed, both used the gender as the variable of interest, however, in the first one, it was evaluated the incidence of COVID-19 infection, and in the second one, the death incidence by COVID-19 among men and women. The search was performed from 7, April 2020 to 20 May 2020. The websites where all information was collected are presented in the supplementary material (Supplementary Table 1).

Generally, the statistics provided by the health ministry of the countries were reported for a sample, thus, the whole country population infected or dead by COVID-19 was estimated proportionally according to the number of infected or deaths reported in the WHO reports in the date of sampling collect.

Men were considered as the treatment group and women the control group. The number of individuals in treatment group (n.e) was represented by the number of men of each country, while the number of individuals in the control group (n.c) corresponded to the number of women in the country. The number of individuals that presented the event in the treatment group (event.e) was the number of infected men, which was estimated in meta-analysis (MA-I) or dead, estimated in meta-analysis (MA-II), the same was set for the control group (female, event.c).

The data collect evaluated the following information: Country; n.e; n.c; sample size; % of male and female of the sample; number of infected and death according to WHO report, estimated male (event.e) and female (event.c) infected and dead by COVID-19.

A total of 38 countries (UK, Italy, Spain, Belgium, Germany, Iran, Canada, Netherlands, Mexico, China, Sweden, Ecuador, Peru, Switzerland, Ireland, Portugal, Indonesia, Romania, Pakistan, Philippines, Japan, Austria, Colombia, Ukraine, Algeria, Denmark, Chile, Dominican Republic, Argentina, Bangladesh, Saudi Arabia, South Africa, Czechia, Finland, Panama, South Korea, Norway and Moldova) were evaluated for MA-I and 41 countries (USA, UK, Italy, France, Spain, Brazil, Belgium, Germany, Iran, Canada, Netherlands, Mexico, China, Sweden, India, Ecuador, Peru, Switzerland, Ireland, Portugal, Indonesia, Romania, Pakistan, Poland, Philippines, Austria, Colombia, Ukraine, Denmark, Chile, Hungary, Dominican Republic, Argentina, Bangladesh, South Africa, Czechia, Finland, Israel, South Korea, Serbia and Norway) for MA-II. It was considered eligible to be included in the analysis those data that presented information of number of infected or deaths by gender in the country.

### Meta-analysis and Meta-regression

The effect size was reported as odds ratio (OR) with respective confidence interval. The random model was chosen because it was observed high heterogeneity of the results among countries. The pooled OR for the random model 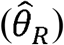 was calculated as follows:

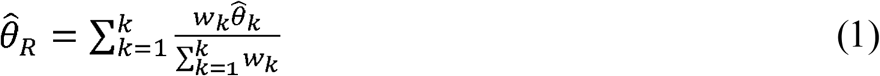

Where, 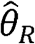 is the weighted average of the 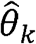 effect sizes from the *k*^*th*^ individual study and *w*_*k*_ is the weight of study *K*, which was calculated by the inverse variance method as 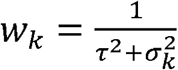. The variance of 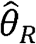 estimator (***τ***^2^) was calculated by using the Sidik-Jonkman estimator and a Hartung-Knapp adjustment:

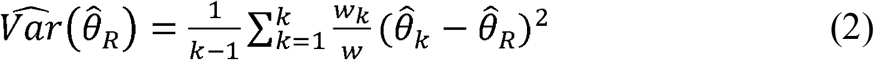

Where *w*_*k*_ was described before and w is 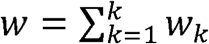. The 1-α confidence interval, with α = 0.05, for the 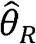 was calculated by:

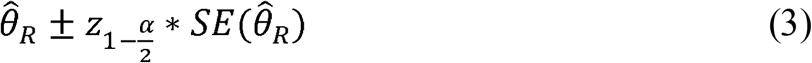

Where 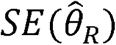 is the standard error of 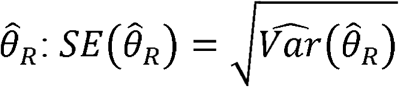 and followed a t-distribution with *k-1* degrees of freedom. The prediction interval was calculated as:

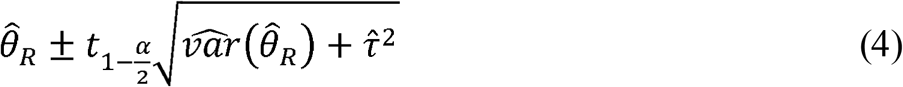

Where 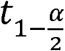 corresponds to the 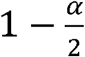 quantile of a t-distribution with *k-2* degrees of freedom. The heterogeneity was accessed by using the Higgins’ and Thompson’s *I*^*2*^ statistic as:

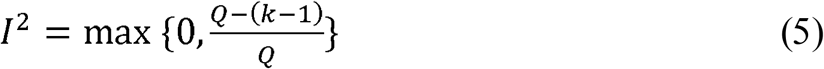

Where Q is the Cochrane Q statistic, obtained as 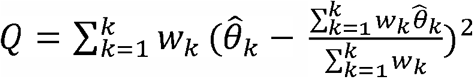, following a qui-squared distribution with *k-1* degrees of freedom. *I*^*2*^ statistic may range from 0% to 100%, *I*^*2*^ = 0% indicates no heterogeneity, from 25% there is low heterogeneity, from 50% medium heterogeneity and from 75% is considered high level heterogeneity. Also, we constructed Labbé plots to access the heterogeneity among countries.

For the meta-regression we used a random mixed effects model because it was assumed that the countries included in this study were a random sample of all countries in the world, i.e., a greater population, that we aim to make inferences. The heterogeneity source was investigated over the variables % of elderly of the country (patients from 60 years old), average age, % of obesity, % of smokers, HDI of the country and number of beds/ 1,000 habitants and average temperature of the last 6 months (from November 2019 to April 2020) of the most affected city, or when this information was not available, it was considered the most populous city of that country. Each variable was evaluated separately. Other variables such as chronic diseases were searched, but the number of countries that presented this information (by gender) was very small, insufficient to perform a meta-analysis. The meta-regression model can be expressed by:

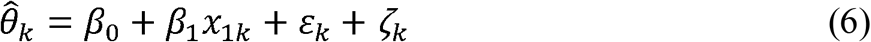

Where 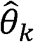 is the effect size of the study *k*, as described before, *β*_0_, *β*_1_ are the unknown regression coefficients, *x*_1*k*_ represents study specific covariables (e.g.: average age), *ε*_*k*_ is the sampling error, which *ε*_*k*_ ∼ *N* (0,1) and *ζ*_*k*_ is the error due to the heterogeneity among studies and *ζ*_*k*_∼ N (0,*τ*^2^). *ε*_*k*_ and *ζ*_*k*_ are assumed to be independent.

Meta-analysis and meta-regression were performed by using *meta* and *metafor* R packages (Balduzzi, Rücker, & Schwarzer, 2019; Viechtbauer, 2010)

## 3. Results

### Meta-analyses

The meta-analysis results for infection and death rate are summarized in Figures 1 and 2. The overall effect of infection rate, 1.05, was close to 1 (*P* = 0.4189), and was not significant, indicating that there was not difference between the infection rate for COVID-19 in men or women. However, for death rate it was observed a greater significant effect size, 1.49 (*P* < 0.0001), meaning a substantial difference between the death rate in both sexes by COVID-19.

**Figure 1.**
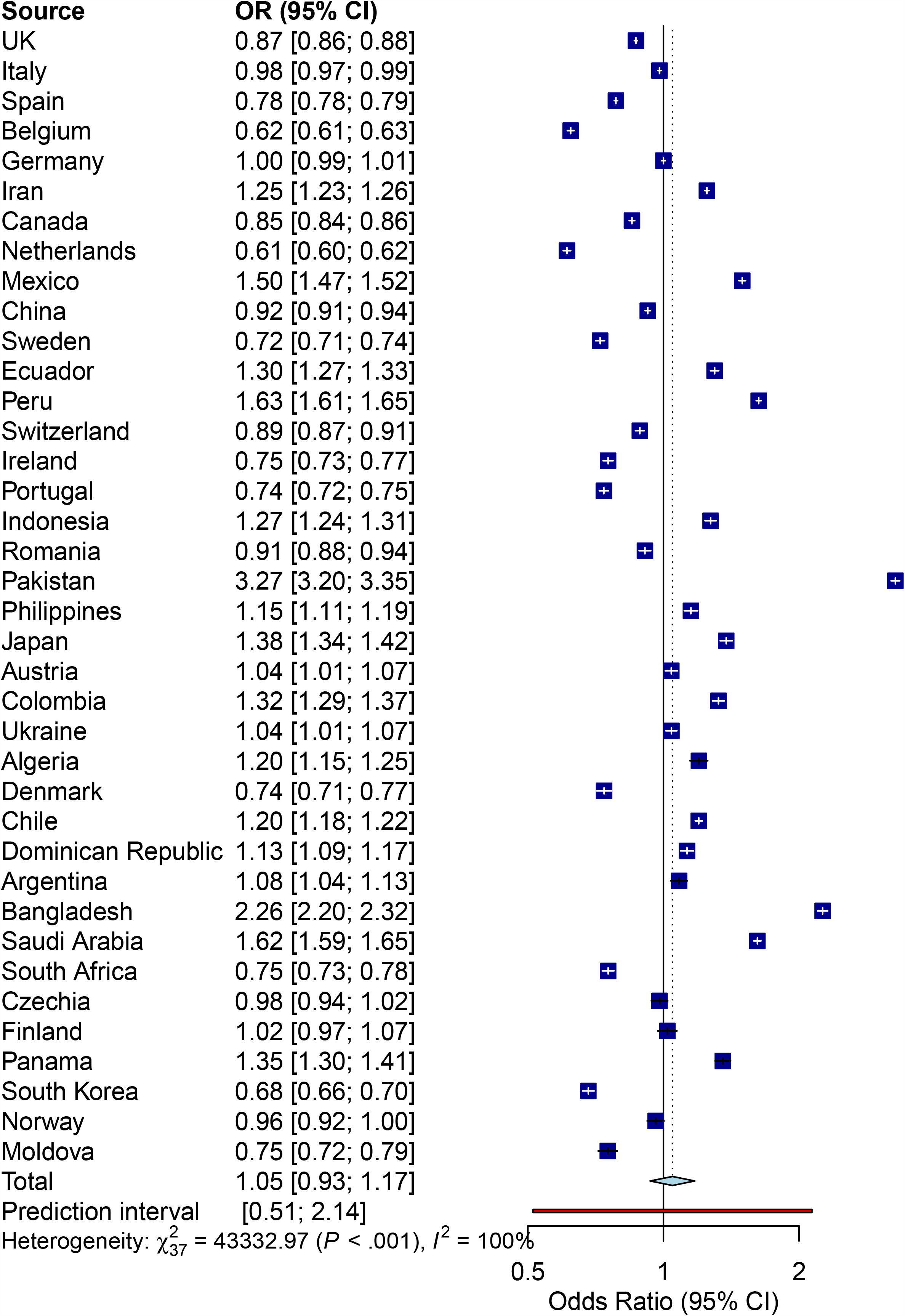
Meta-analysis of infection rate by COVID-19 by gender in the top 50 countries with highest death rates.

**Figure 2.**
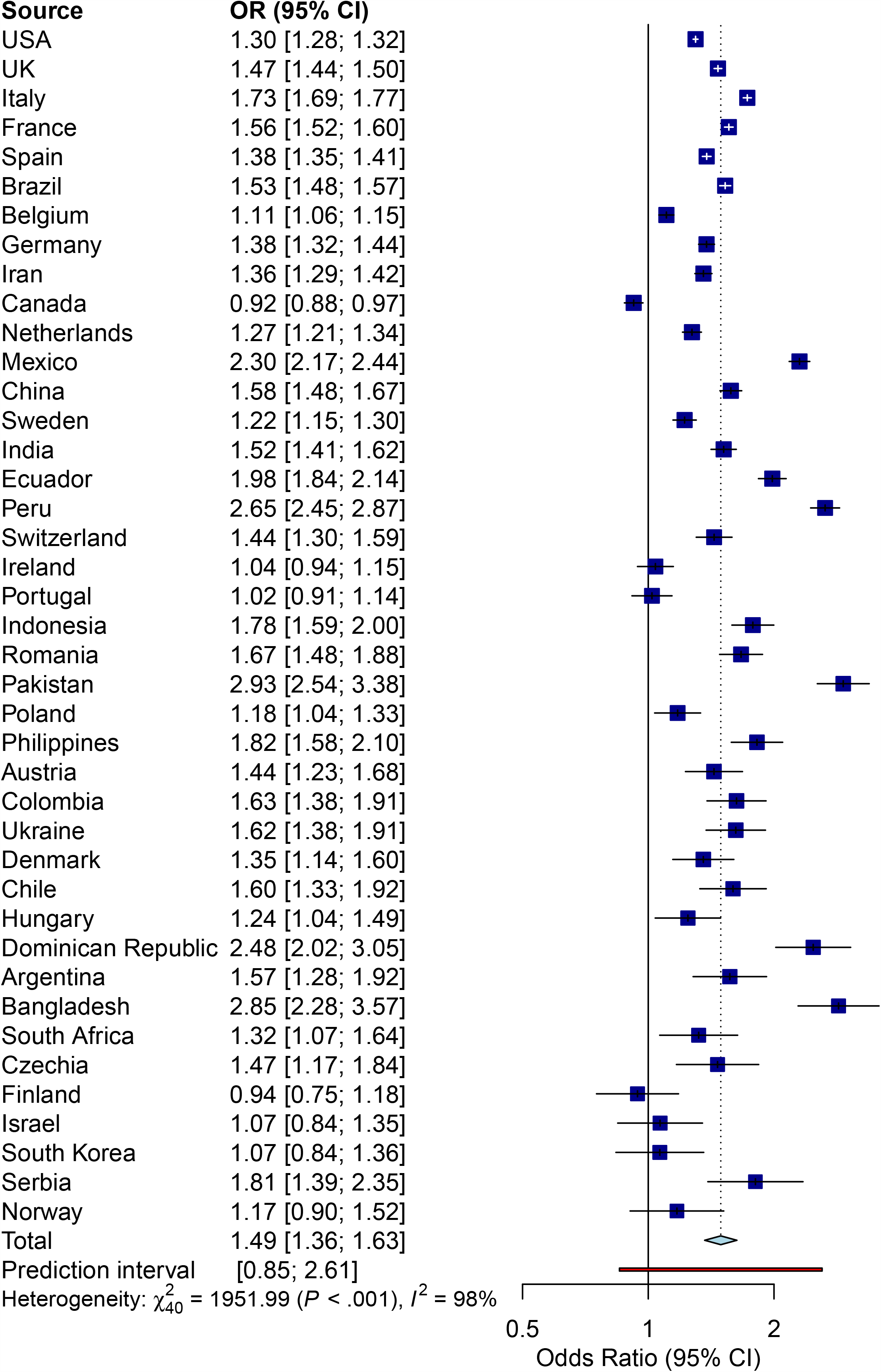
Meta-analysis of death rate by COVID-19 by gender in the top 50 countries with highest death rates.

Prediction interval ranged from 0.51 to 2.14 for infection rate and from 0.85 to 2.61 for death rate, which means that we do not have a great confidence about the infection or death rate would be always higher in man than in woman, however this uncertain is smaller for death rate than for infection rate.

### Heterogeneity among countries and meta-regression

As expected, heterogeneities of both meta-analyses were high (> 75%). Differences in infection and death rates among countries are common as different sources of variability occur around the word.

Figures 3 and 4 present the Labbé plots of both meta-analyses. By the visual inspection of these figures, we observed that there was a little difference in the incidence rate of infection among control (women) and treatment (men) groups and a larger difference among them for death rate (because the red line was more far from the blue line in Figure 4), however both plots showed the stronger trend of men for infection and death (the red line follow a trend to the left and upper side), indicating a negative effect of the treatment group compared to the control group. Italy, Spain and UK presented high incidence rate (proportional to the country population size) for death in comparison with the other countries.

**Figure 3.**
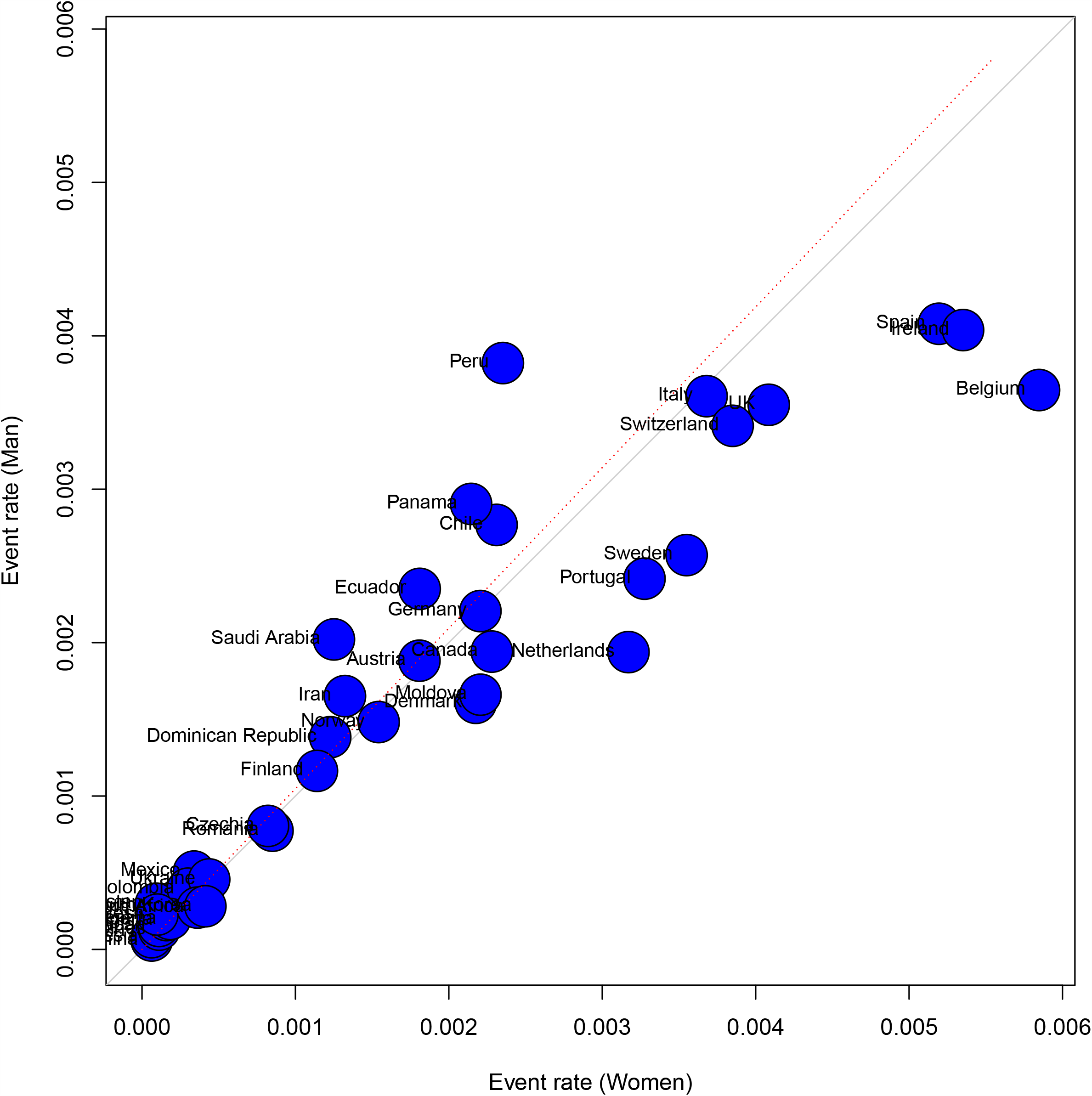
Labbé Plot of infection rate by COVID-19. The blue line represents no effect. The dotted red line indicates the pooled odds ratio effects. Blue circles denote each individual study and the size of the circle reflects the study weight in the meta-analysis.

**Figure 4.**
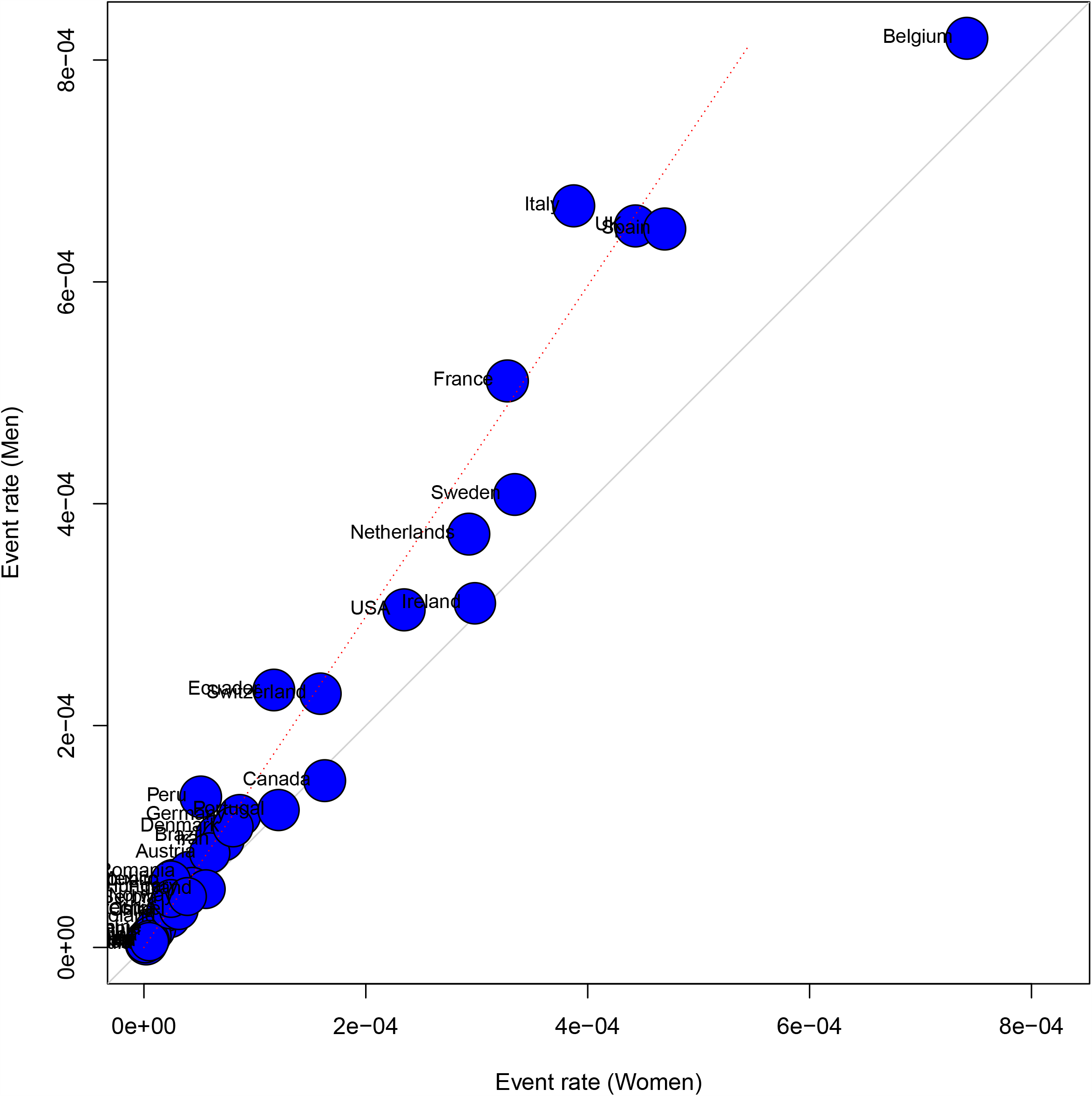
Labbé plot for death rate by COVID-19. The blue line represents no effect. The dotted red line indicates the pooled odds ratio effects. Blue circles denote each individual study and the size of the circle reflects the study weight in the meta-analysis.

Another information that we can extract from Figure 4 is about the heterogeneity of four studies, Italy, Spain, UK and Belgium were located more far than the other countries in the death rate Labbé plot. This result suggest that they are contributing to the heterogeneity across countries. Italy, UK and Belgium are contributing to the heterogeneity of the effect sizes, because their OR deviated a little from the overall effect, but Spain contributed to the heterogeneity of the event rates, because the number of deaths was one of the highest considering the proportionality of the total population size. Regarding Figure 3, all countries that are more far from the blue line (Spain, Ireland, Belgium and Peru) were considered outliers by their OR.

Meta-regressions were conducted to try to explain the variability source among countries. Different variables were evaluated: elderly percentage of the country, average age, obesity percentage, smoker percentage, average temperature of the last 6 months in the most affected city, number of hospital beds for each 10,000 hab. and HDI. These variables were evaluated according to the proportion of heterogeneity that they were able to capture, i.e., highest R^2^ and p-value < 0.05.

The percentage of heterogeneity take into account by each variable is presented in Table 1. HDI was the most important parameter explaining the most part of variability among countries for both infection and death rates, and was higher for death rate. Other parameters were also significant for both rates, besides HDI: the average age of the country and temperature (p < 0.001). For death rate, number of hospital beds available per 1,000 habitants and obesity percentage presented a smaller significance (p < 0.05).

**Table 1.** Percentage of heterogeneity take into account by each parameter evaluated (R^2^) and their significance.

| eta-regression<br>parameters | Trait |  |
| --- | --- | --- |
| | $R^2$ Infection rate | $R^2$ Death rate |
| A. Age<sup>1</sup> | 40.89<sup>a</sup> | 40.27<sup>a</sup> |
| A. Temperature<sup>2</sup> | 30.06<sup>a</sup> | 39.26<sup>a</sup> |
| HDI | 42.31<sup>a</sup> | 54.21<sup>a</sup> |
| n. beds | 10.51<sup>d</sup> | 13.85<sup>c</sup> |
| % elderly | 7.29<sup>ns</sup> | 1.26<sup>ns</sup> |
| % obesity | 4.57<sup>ns</sup> | 10.81<sup>c</sup> |
| % smokers | 0.38<sup>ns</sup> | 0.80<sup>ns</sup> |

<sup>1</sup>A. age = Average age; <sup>2</sup> A. Temperature = Average temperature of 6 months (Nov-Apr); 3= upper scripts indicate different significance levels (a = $p < 0.001$ ; b = $p < 0.01$ ; c = $p < 0.05$ ; d = $p < 0.1$ ; ns = no significance).

The top 4 significant variables were closely evaluated for their significance and possible interactions in a multi-model inference analysis for infection and death rates, but no significant interactions were observed (data not shown).

Curiously, countries with smallest HDI (Pakistan and Bangladesh) were the same that presented highest difference among sexes, for both infection and death rates. In these countries the rate of infection and death was much greater for man than for woman. By other side, countries with highest HDI (Norway and Switzerland) presented rates more egalitarian between sexes. HDI is an important index that measure the degree of human development across countries, and this index seems to be also reflecting the capacity of the countries to prevent and save lives against COVID-19, possibly because it reflects the capacity of people of social isolation, keeping themselves safe.

## 4. Discussion

This meta-analysis study included the information infection and death rates among sexes of the top 50 countries with the highest number of deaths until 20^th^ May, provided by the WHO and individual governments. The results pointed that there was no significative difference in infection rate for male and female by COVID-19. However, significative difference was observed for death rate, showing that men are more susceptible to death than women. Heterogeneity was observed among countries for both meta-analysis and the factors that explained more were: HDI, average age, temperature, but the number of hospital beds was also important for death rate.

Women are less susceptible to viral infections than men because of a series of factors such as differences in innate immunity, hormones and sex chromosomes. Women present two X chromosomes, which reinforce the immune system activity, even though one of them is inactive. This chromosome presents genes related to the immune system that play important roles in fighting against virus and general inflammations. This fact could be one of the possible reasons that explains why the death rate was generally higher across countries in men compared to women, even though there was a little overall difference for most of the countries in the infection rate between sex (Conti & Younes, 2020). According to Jin et al. (2020), that studied the gender differences in 37 patients with COVID-19 in China, men and women have the same prevalence but men with COVID-19 are more prone for worse outcomes and death, independently of age.

Among the evaluated countries, Pakistan and Bangladesh presented the highest odds ratio, indicating that in these countries, men were more infected and died more by COVID-19 than women. By other side, the smallest odds ratios were found for Belgium and Netherlands, for infection and Canada and Finland for death.

Average age was another factor important in both analyses. The age of patient is an important factor to determine a trend to respiratory infections. Meyer (2004) explains that elderly present a slower immune response and frequently present some organ disfunction related to the age, which also is associated with the presence of chronic diseases. Furthermore, the mortality and morbidity in this age are often higher than in young ages. Cromer et al. (2014) investigated the influenza burden by age in United Kingdom and verified that the majority of deaths were elderly people with co-morbidities.

Temperature was also significant for infection and death and it is another factor that may affect the health of respiratory tract. Mäkinen et al. (2009) concluded that individuals exposed to low temperatures were more prone to present common cold, pharyngitis and lower respiratory tract infections. An interesting association between cold temperatures and H1N1 influenza virus was found in Netherlands (Kunst, Looman, & Mackenbach, 1993). In a period of eight years they observed the behavior of death rate during hot and cold periods controlling different variables. When influenza was controlled, the death rate during the cold decreased 34%, while heat-rate was 23%. Wang et al. (2020) also tested the influence of temperature and humidity on number of cases in China and US, and they concluded that there is some influence of these climate factors in COVID-19 incidence, however they highlighted that public politics encouraging social distance would be more effective than those climate factors isolated.

Number of hospital beds in a country is a parameter that may reflect the capacity of a country to take care of COVID-19 patients, giving the necessary medical care that they will need, and is also kind of related to HDI. The number of beds range widely from a city to other, and a more precise evaluation could take into account for example, the city with highest number of cases or deaths, however this information generally is not accessible.

Obesity percentage have also been included as a morbidity predisposing patients to severe COVID-19 cases. Meta-regression results showed some significance of this factor to explain the heterogeneity across countries. According to Dietz & Santos□Burgoa (2020), in California from patients with influenza that presented obesity, 66% presented other kind of underlying diseases, as respiratory chronic diseases, heart problems or diabetes. The obese patients presented their pulmonary function compromised and generally present increased inflammatory cytokines.

Besides smokers percentual was not significative, the countries with highest odds ratio, Pakistan and Bangladesh presented high rate of man smokers, much more than women (gender difference of 41.8% for Pakistan and 34% for Bangladesh). Tobacco consumption is related to an increasing of respiratory tract infection caused by influenza or pneumonia infections. The risk of influenza infection in smokers was 5 times higher compared to non-smokers (Lawrence et al., 2019). Furthermore, the age is also related with the infection risk of respiratory tract. Liu et al. (2013) concluded in a trial study that Chinese patients infected with avian influenza A presented higher probability to death by increasing age, smoking, previous known diseases or using chronic drugs.

Other morbidities are also important to complicate COVID-19 cases, such as pre-existing diseases, as diabetes, cardiopathies, renal problems, etc. However, there are limiting factors to evaluate their impact because of no information about this is available publicly for most countries. Other factors such as the number of hospitalizations and patients in intensive care unit by COVID-19 would also be interesting to analyze but it is also difficult to obtain this information. Furthermore, these results are regarding to an ongoing outbreak, in which the numbers change constantly, and the information is not available for all countries.

## 5. Conclusion

Men are dying more than women. No significant difference was observed for infection rate between genders. HDI was the most important factor affecting both infection and death rates. Countries with highest HDI present less difference between sexes. This could be an evidence of the importance of public politics to promote the general well-being of the population equally.

## Data Availability

The data is publicly available in the official bodies of the countries studied.

https://globalhealth5050.org/covid19/

https://data.humdata.org/

https://data.oecd.org/healtheqt/hospital-beds.htm

## 6. Declaration of Competing Interest

The authors declare that there is no conflict of interests.

## 7. Acknowledgments

This work was supported by the national post-doctoral program of CAPES – Higher Education Improvement Coordination agency, Grant [number 88887.355147/2019-00].

## Notes

### Competing Interest Statement

The authors have declared no competing interest.

